# Functional connectivity in preterm infants with intraventricular hemorrhage using fNIRS

**DOI:** 10.1101/2024.02.27.24303403

**Authors:** Lilian M. N. Kebaya, Lingkai Tang, Talal Altamimi, Alexandra Kowalczyk, Melab Musabi, Sriya Roychaudhuri, Homa Vahidi, Paige Meyerink, Paula Camila Mayorga, Sandrine de Ribaupierre, Soume Bhattacharya, Leandro Tristao Abi Ramia de Moraes, Michael T. Jurkiewicz, Keith St. Lawrence, Emma G. Duerden

**Affiliations:** Neonatal-Perinatal Medicine, Schulich Faculty of Medicine and Dentistry, Western University, London, Ontario, Canada; Neuroscience, Schulich Faculty of Medicine and Dentistry, Western University, London, Ontario, Canada; Biomedical Engineering, Faculty of Engineering, Western University, London, Ontario, Canada; Clinical Neurological Sciences, Schulich Faculty of Medicine and Dentistry, Western University, London, Ontario, Canada; Medical Imaging, London Health Sciences Centre, Western University, London, Ontario, Canada; Medical Biophysics, Schulich Faculty of Medicine and Dentistry, Western University, London, Ontario, Canada; Applied Psychology, Faculty of Education, Western University, London, Ontario, Canada

**Author notes:** **Corresponding author:** Address correspondence to: Emma G. Duerden, PhD Applied Psychology, Faculty of Education 1137 Western Rd London, Ontario N6G 1G7. These authors contributed equally to this work (Lilian M. N. Kebaya, Lingkai Tang).

**Keywords:** intraventricular hemorrhage, functional connectivity, functional near-infrared spectroscopy

## Abstract

**Introduction:** Intraventricular hemorrhage (IVH) is a common neurological complication following very preterm birth. Resting-state functional connectivity (RSFC) using functional magnetic resonance imaging (fMRI) is associated with injury severity; yet fMRI is impractical for use in intensive care settings. Sensitive bedside neuroimaging biomarkers are needed to characterize injury patterns. Functional near-infrared spectroscopy (fNIRS) measures RSFC through cerebral hemodynamics and has greater accessibility. We aimed to determine comparability of RSFC in preterm infants with IVH using fNIRS and fMRI at term equivalent age (TEA), then examine fNIRS connectivity with the severity of IVH.

**Methods:** Very preterm born infants with IVH were scanned with both modalities at rest at TEA (postmenstrual age=37±0.92 weeks). Connectivity maps of IVH infants were compared between fNIRS and fMRI with the Euclidean and Jaccard distances. The severity of IVH in relation to fNIRS RSFC strength was examined using generalized linear models.

**Results:** fNIRS and fMRI RSFC maps showed good correspondence. At TEA, connectivity strength was significantly lower in healthy newborns (p-value = 0.023) and preterm infants with mild IVH (p-value = 0.026) compared to infants with moderate/severe IVH.

**Conclusion:** fNIRS has potential to be a new tool for assessing brain injury and monitoring cerebral hemodynamics and a promising marker for IVH severity in very preterm born infants.

**Highlights:**

- There is no previous study combining fNIRS and fMRI focused on preterm neonates with IVH.
- This is the first study associating functional connectivity strength yielded from fNIRS with severity of IVH.
- Good correspondence of functional connectivity was shown between fNIRS and fMRI.
- Preterm neonates showed increased functional connectivity strength compared to healthy term born neonates. This can potentially be a marker for clinical assessment of severity of IVH.
- fNIRS was demonstrated to have potential as a new bedside neuromonitoring tool for assessing early brain injuries of neonates.

## Introduction

Germinal matrix-intraventricular hemorrhage (GMH-IVH) continues to be a major morbidity amongst neonates born premature prior to 32 weeks of gestation [1]–[3]. Based on the severity, GMH-IVH is classified into four grades: grade 1, hemorrhage confined to the germinal matrix; grade 2, occupying <50% of the ventricle; grade 3, distending and occupying >50% of the ventricle, and grade 4 – IVH with intraparenchymal hemorrhage [4]. Severe GMH-IVH (grade III/IV) directly or its consequence such as post-hemorrhagic ventricular dilatation (PHVD) may lead to injury to the developing periventricular white matter that can be seen on conventional imaging at term equivalent age (TEA) [5]. Furthermore, some of these effects persist later in life, impacting language, cognitive, behavioural, and motor domains [6].

More recently, the use of resting-state functional connectivity (RSFC) revealed with functional magnetic resonance imaging (fMRI) has been employed to gain a better understanding of brain injury and the impact on brain function, indicating this can be a promising biomarker [7]–[10]. RSFC, through investigating the correlations in spontaneous blood oxygenation level-dependent (BOLD) signal fluctuations, has provided new perspectives for studying brain injury in preterm infants [8],[10],[11]. fMRI-based RSFC in neonates with perinatal brain injury was predictive of motor skills at 8 months of age [12]. Yet, the clinical utility of functional MRI is limited due to its accessibility and may be impractical for use with the sickest neonates who have severe IVH who cannot undergo transport to MRI.

Functional connectivity determined from fMRI shows signal alterations based on BOLD signal and is reflective of deoxygenated hemoglobin (Hbr) [13]. Functional near infrared spectroscopy (fNIRS) is a non-invasive and light-based brain imaging technique that can be used to map functional connectivity at the bedside. fNIRS exploits the process of neurovascular coupling and measures the absorption of near-infrared light by hemoglobin [9]. Hence, oxygenation of the cerebral cortex is derived as an indirect measurement of neural activity. Both indices reflective of cerebral oxygenation, Hbr and oxygenated hemoglobin (HbO) along with total Hb can be estimated using fNIRS. fNIRS is an extremely convenient neuromonitoring tool, that can be acquired at the bedside, within a short time period with minimal inconvenience to fragile neonates.

GMH-IVH, regardless of grade, has been associated with disrupted functional connectivity in neonates [14]–[16]. RSFC has been demonstrated to be associated with ventricular volumes in very preterm born neonates with PHVD [16], indicating that this method can reliably be used at the bedside in the neonatal intensive care unit (NICU). Given the strong need for bedside tools to monitor injury patterns in very preterm born neonates and evidence suggesting that fNIRS based RSFC is comparable to fMRI in adults [17], we sought to compare the RSFC maps acquired using fNIRS and fMRI in very preterm neonates with IVH who were assessed at TEA. We also aimed to investigate whether fNIRS-based RSFC in preterm infants would be associated with the severity of IVH, compared to healthy term-born neonates. Our overall hypothesis was that fNIRS-based measures of RSFC would be comparable to that obtained with fMRI and that severity of injury would be associated with changes in RSFC patterns.

## Methods

### Participants

This was a prospective cohort study. Study participants were recruited from the NICU at the London Health Sciences Centre (LHSC), London, Canada between January 2020, and December 2022. Preterm neonates were eligible for inclusion based on the following criteria: ≤ 32 weeks’ gestational age (GA), born at, or referred to NICU, and admitted with a diagnosis of GMH-IVH, made by the most responsible physician on the infant’s first routine cranial ultrasound. Exclusion criteria were the following: major anomalies of the brain or other organs, congenital infections, intrauterine growth restriction, metabolic disorder, and ultrasound evidence of a large parenchymal haemorrhagic infarction. Term-born infants with no reported brain injury were recruited as healthy controls. Participants were recruited from the LHSC Mother baby Care Unit (MBCU). Inclusion criteria were birth >36 weeks’ GA, born at LHSC, and admitted to MBCU. Exclusion criteria were the following: congenital malformation or syndrome, antenatal infections, antenatal exposure to illicit drugs, small for gestational age and intrauterine growth restriction.

### Clinical variables

The neonatal charts were reviewed by Neonatal-Perinatal Medicine Fellows (LMNK, TA, SR, MM), Paediatric Resident (AK), Neuroscience Graduate Student (HV) or NICU Nurses (PM, CM) for demographic and clinical characteristics. The following postnatal events were included: days of mechanical ventilation, bronchopulmonary dysplasia, patent ductus arteriosus requiring treatment, days of parenteral nutrition, culture proven sepsis, necrotizing enterocolitis.

### MRI acquisition & image analysis

Anatomical and functional MRI images were acquired on a 1.5 T GE scanner at LHSC. Each infant underwent a clinical MRI scan consisting of a whole-brain T1-weighted structural image (TR=8.4–11.5 ms [depending on clinical requirements], TE=4.2 ms, flip angle=12/25°, matrix size 512 × 512, 99–268 slices, voxel size typically 0.39 × 0.39 × 0.5 mm (0.31 × 31 × 5 to 0.43 × 0.43 × 0.6 for some infants), and a T2-weighted structural image (TR=3517–9832 ms, TE=7.3–8.4 ms, flip angle = 90/160°, matrix size 256 × 256, 19–60 slices, 0.7 × 0.7 × 2–5 mm voxel resolution). Blood oxygen level–dependent (BOLD) fMRI data were acquired using an echo planar imaging sequence to examine resting-state functional connectivity (TR=3000ms, TE=50 ms, flip angle=70°, matrix size 64 × 64, 39 slices, voxel size 3 x 3 x 3 mm, total volumes 35).

Preprocessing of fMRI images was conducted with FMRIB Software Library (https://fsl.fmrib.ox.ac.uk/fsl/fslwiki/FSL). The pipeline included brain extraction, motion correction, spatial smoothing (full width at half maximum = 5 mm), band-pass filtering (0.01-0.2 Hz) and registration to a neonatal atlas [18]. Average BOLD sequences were extracted from frontal, parietal, temporal and occipital lobes of both hemispheres, and then correlated to build an 8-by-8 lobe-wise RSFC map for each infant.

### Brain injury characterization

A Neuroradiology Fellow (LTARdM) scored the T1-weighted anatomical images for brain injury severity. These were verified by a Paediatric Neuroradiologist (MTJ). IVH was graded (mild=1-2, and moderate-severe=3-4) using Papile’s method[4].

### fNIRS acquisition & analysis

All participants (preterm infants with IVH and healthy newborns) were scanned with a NIRSport2 (NIRx, Berlin, Germany) unit with two emission wavelengths (760 and 850 nm). We used an 8-by-8 set up with 20 channels covering the whole brain, and a sampling rate of 10 Hz (Fig 1.A). For each hemisphere, there were 4 channels on the temporal, 2 on the parietal, frontal and occipital lobes. Scans were recorded for a minimum of 6 minutes during rest or natural sleep with the participant either lying in the incubator, cot, or caregiver’s arms.

**Figure 1.**
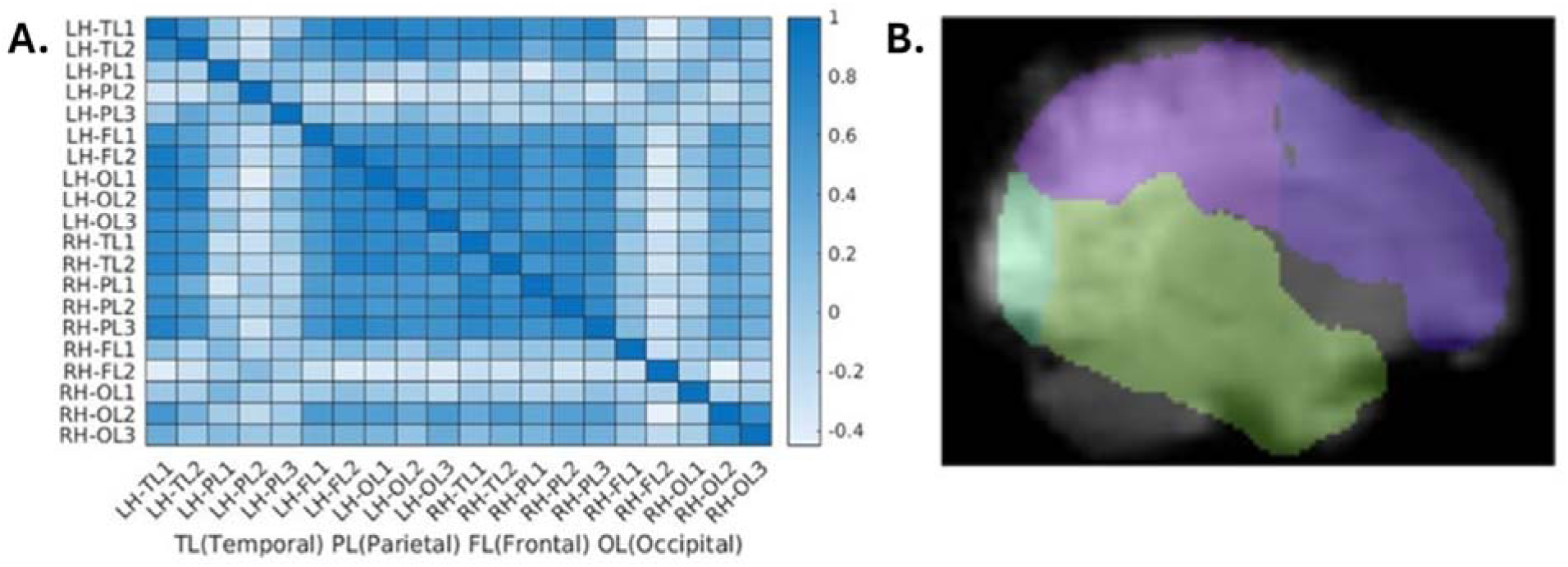
A. Connectivity matrix for the signals extracted from the fNIRS channels in the left (LH) and right (RH) hemispheres from the temporal, parietal, frontal and occipital lobes. B. Lobe-based analysis of the fMRI data. A hemisphere is sectioned into temporal, parietal, frontal and occipital lobes.

After data acquisition, for fNIRS recording of each infant, visual inspection was done to select a 6-minute segment with fewer motion artifacts and less background noise. A preprocessing pipeline, built within Homer 3 software, including spline interpolation for motion correction [19], Savitzky–Golay filtering with frame size of 10 [20], band-pass filtering of 0.01-0.1 Hz and modified Beer-Lamber law to convert optical density to concentration change of HbO and Hbr, respectively [21]. Pearson correlation was used to calculate connectivity. To compare RSFC maps between fNIRS and fMRI, channels corresponding to one lobe were averaged then correlated with other cortices to create an 8-by-8 lobe-wise connectivity maps for HbO and Hbr, respectively (Fig 1.C). For lobe-wise maps of both fNIRS and fMRI, nodes were lobes and edges were weighted by Pearson correlation coefficients between sequences of the two lobes. 20-by-20 channel-wise RSFC maps were also calculated for fNIRS (Fig 1.B), yet in this map, nodes were fNIRS channels.

### Statistical analysis

Statistical analyses were performed using a combination of Matlab (R2020b, Natick, Massachusetts: The MathWorks Inc) and Statistical Package for the Social Sciences (v.29, Chicago, IL).

To address our first aim, whether the RSFC maps rendered from the fNIRS and fMRI modalities were comparable, we calculated the Euclidean distances and Jaccard distances between the two lobe-wise maps at various levels of sparsity for both weighted and binarized maps [22]. The sparsity was the percentage of non-zero connections in a RSFC map after all negative-weighted and some low-weighted connections were set to zero. Weighted maps were obtained in this way. Binarized maps were then calculated with non-zero connections set to 1. Given maps *G_fMRI_*(*V*, *E_fMRI_*) and *G_fNIRS_*(*V*, *E_fNIRS_* obtained from the two modalities, where *V* and *E* denote node and connection set, respectively, and their adjacency matrices 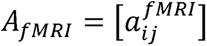 and 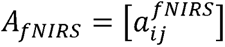, respectively, where and are nodes, Euclidean distance between two maps was defined as

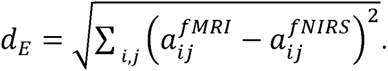

Note that *A* is the average over all subjects’ individual adjacency matrices. Jaccard distance for weighted maps was defined as

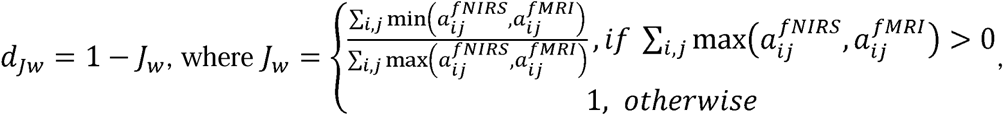

while for binarized maps, Jaccard distance was defined as

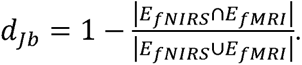

One level of sparsity yielding the most similarity between two modalities was picked for following analyses. Based on this specific sparsity, we also calculated similarity maps for weighted and binarized RSFC maps. Similarity map was defined as

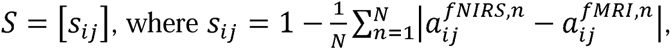

where 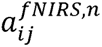 and 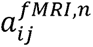 are entries of induvial adjacency matrices of, *n^th^* subject from a total of *N*. Note that for weighted RSFC maps, *S_ij_* was an index based on Euclidean distance, while for binarized maps, *S_ij_* equals to the percentage of subjects sharing the connection.

We addressed our second aim, concerning whether fNIRS-based RSFC would predict IVH severity in very preterm born neonates compared to healthy newborns, in generalized linear models. In the first model, the summation of weighted connectivity values of the whole brain for Hbr were entered as the dependent variable, with IVH grade (none [healthy newborns], mild, moderate-severe) as a factor, adjusting for biological sex, GA, and postmenstrual age (PMA) at scan. In the second model, HbO connectivity values were entered as the dependent variable, using the identical independent variables as in the first model. As we had a single hypothesis regarding the association of IVH grade compared to healthy newborns, the alpha level was set at p=0.05.

## Results

### Participants

Sixteen very preterm born neonates with GMH-IVH were enrolled (GA at birth = 26.28±2.82 weeks). All preterm infants underwent MRI scanning at TEA (GA at scan = 37.04±0.96 weeks). In eleven participants, fNIRS was also acquired. A total of 15 term-born infants were recruited from the LHSC MBCU with mean birth GA of 38.92±1.30 weeks and fNIRS scans performed within 48 hours of life. None of the healthy newborns underwent MRI.

### Comparing fNIRS and fMRI

Lobe-wise connectivity maps were obtained from fNIRS for both HbO and Hbr. The fNIRS maps were then compared against fMRI RSFC maps at various levels of sparsity using metrices of Euclidean and Jaccard distances. Only large- and positive-weighted connections were kept when achieving for a certain level of sparsity. Sparsity ranged from 0.2 to 0.4. At 32% of sparsity, for both weighted and binarized maps, for both HbO and Hbr maps, least Euclidean and Jaccard distances were achieved (i.e., the fNIRS and fMRI yielded most similar RSFC maps, Fig. 2). Therefore, in the subsequent analyses, this level of sparsity was applied.

**Figure 2.**
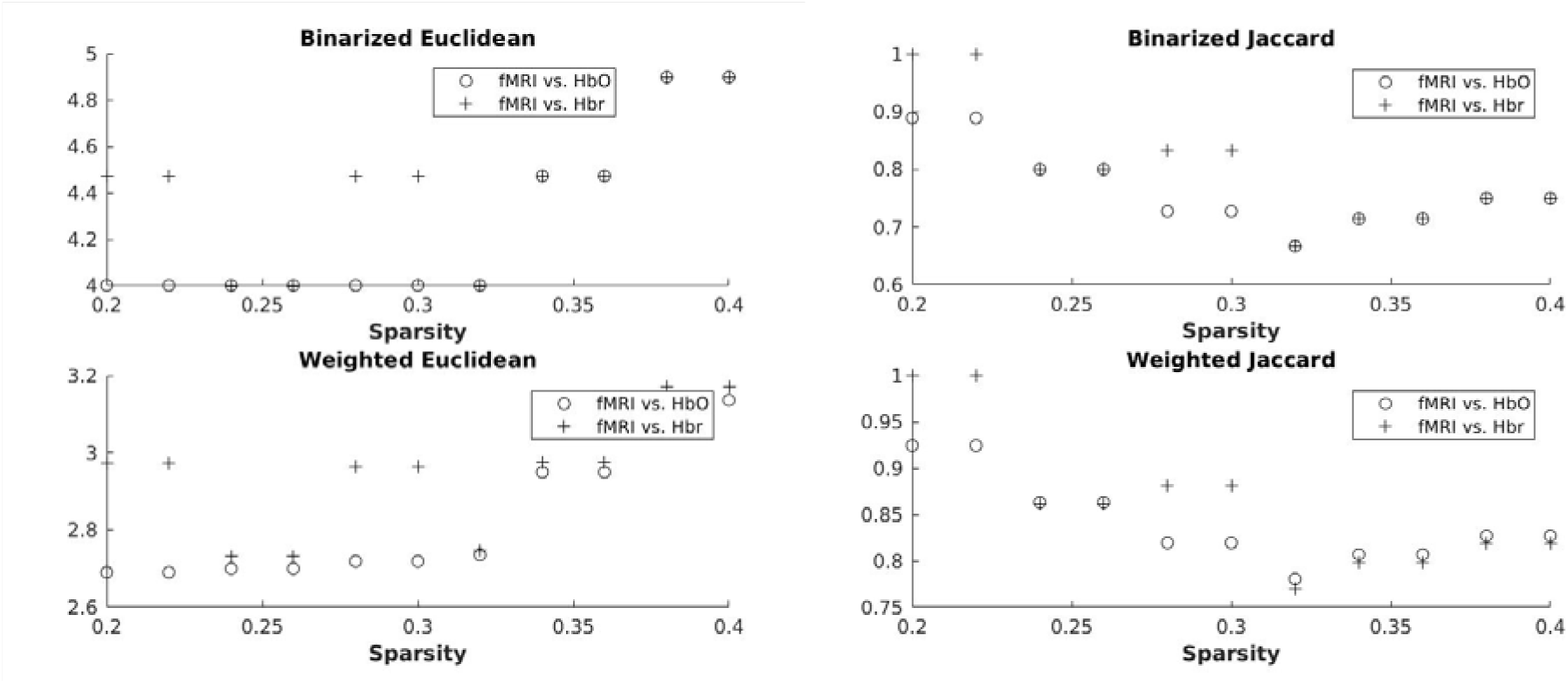
Similarity of RSFC maps between fNIRS and fMRI with sparsity, measured by Euclidean distance on binarized maps (upper left), Euclidean distance on weighted maps (lower left), Jaccard distance on binarized maps (upper right), and Jaccard distance on weighted maps (lower right).

Based on the sparsity of 32%, the similarity maps were calculated comparing every connection between fNIRS and fMRI. Similarity maps of HbO against fMRI and Hbr against fMRI had good correspondence for both weighted and binarized cases (Fig. 3). This is largely a reflection of the anti-correlation between the two chromophores, yielding similar RSFC maps. Also, most connections demonstrating high correspondence were interhemispheric.

**Figure 3.**
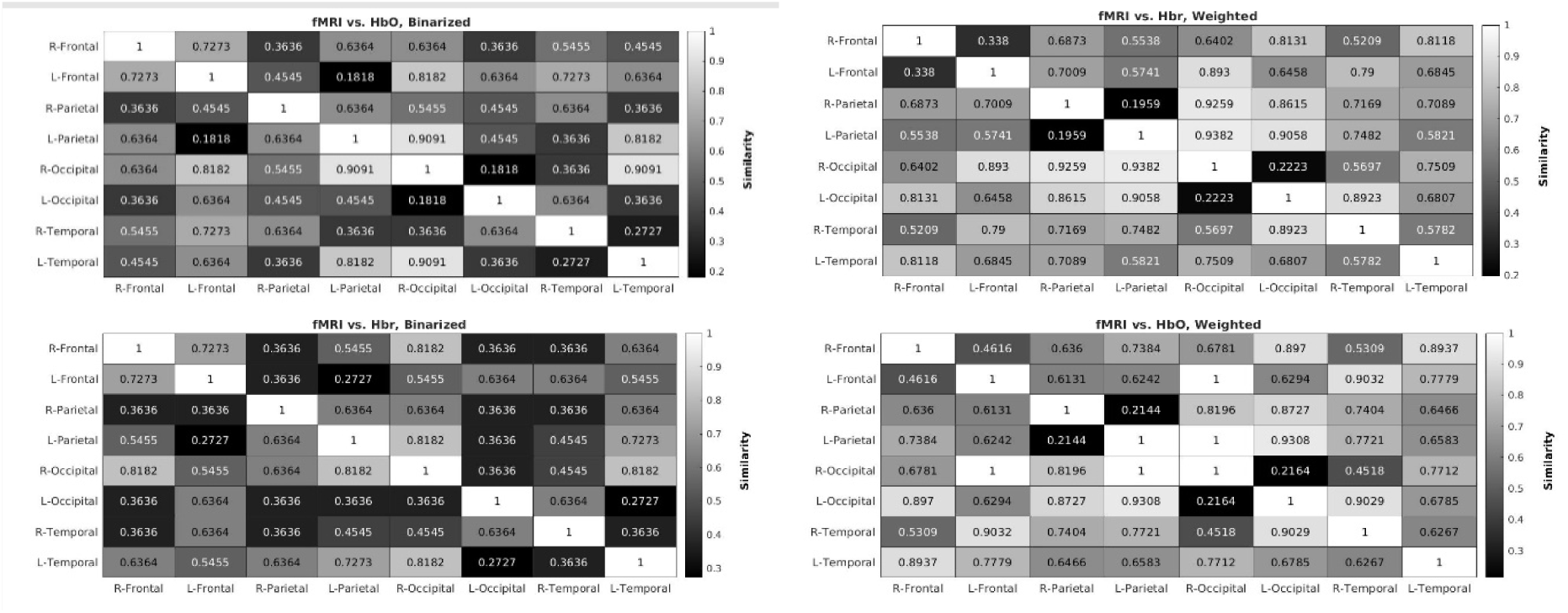
Lobe-wise similarity maps at sparsity of 32%. Higher values reflect greater similarity between the two modalities. Average values among channels are 0.5390 (upper left), 0.5260 (lower left), 0.7031 (upper right) and 0.6761 (lower right).

### Connectivity and severity of IVH

In a generalized linear model, the functional connectivity within the cortical network was examined in relation to IVH severity (none [healthy newborns], mild IVH, moderate/severe IVH). The weighted connectivity values for Hbr were significantly lower in healthy newborns (B= -53.5, 95%CI -94.61 - -12.39, p=0.011, Table 2, Fig 4), and preterm neonates with mild IVH (B=-24.7, 95%, CI -42.7- -6.7, p=0.007) compared to preterms with moderate/severe IVH adjusting for birth GA, sex, and PMA at scan. No significant differences in Hbr connectivity values were evident between the healthy newborns and very preterm born neonates with mild IVH (p=0.5, Bonferroni corrected for multiple comparisons). Similar results were found when examining HbO in a separate GLM, whereby healthy newborns (B=-42.6, 95%, CI -79.1- -6.0, p=0.023, Table 2, Fig 4) and very preterm neonates with mild IVH (B=-18.2, 95%, CI -34.3- - 2.2, p=0.026) compared to very preterm born neonates with moderate/severe IVH. No differences were evident between the HbO sparsity values for healthy newborns and neonates with mild IVH (p=0.6, Bonferroni corrected for multiple comparisons).

**Figure 4.**
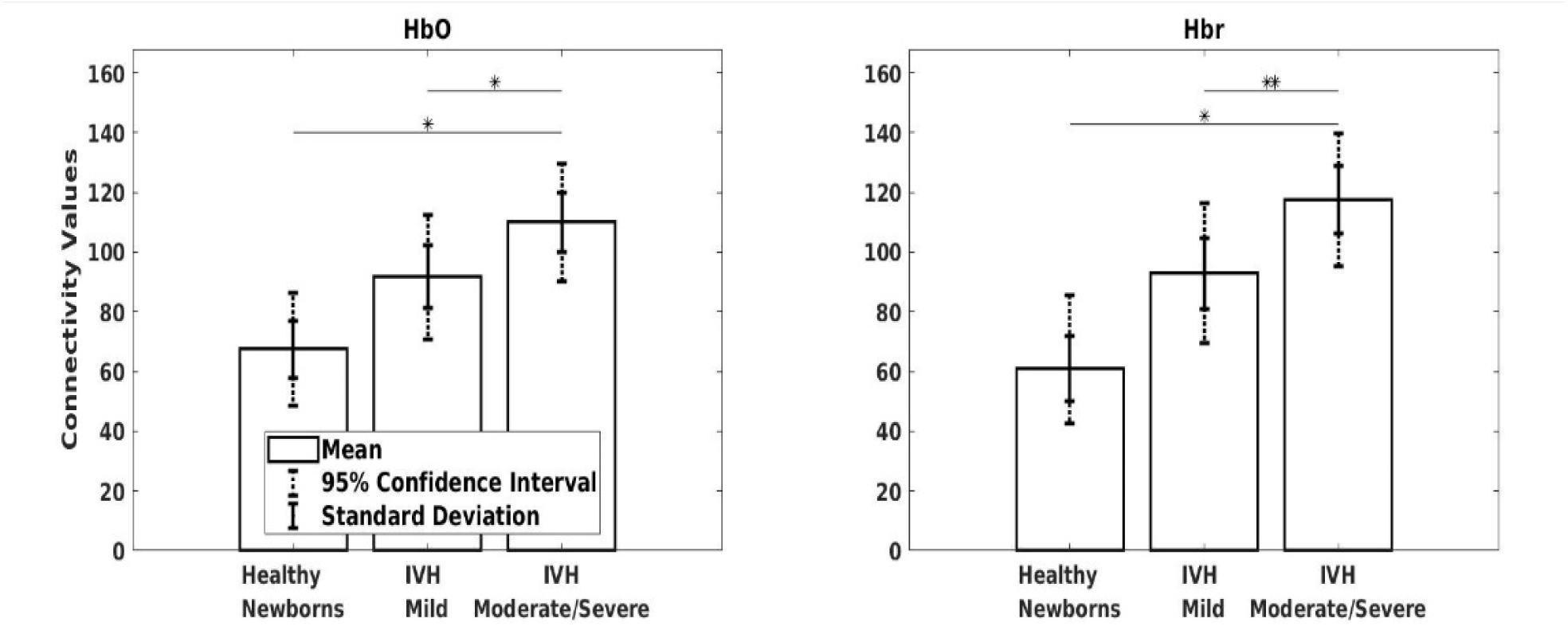
Very preterm born neonates scanned at term-equivalent age showed increased HbO (left) and Hbr (right) connectivity values relative to healthy newborns and neonates with mild IVH. **p<0.01, * p<0.05.

**Table 1.** Participant demographics.

|  | Neonates with IVH | Healthy Newborns |
| --- | --- | --- |
| *n* | 16 | 15 |
| Male sex | 12 | 6 |
| Birth GA (SD) | 26.1(2.8) | 38.9(1.3) |
| PMA at scan (SD) | 36.9(0.9) | 38.9(1.3) |
| IVH severity |  |  |
| mild | 7 | - |
| moderate/severe | 9 | - |

Values represents counts for categorical variables and means for continuous variables.

**Table 2.**
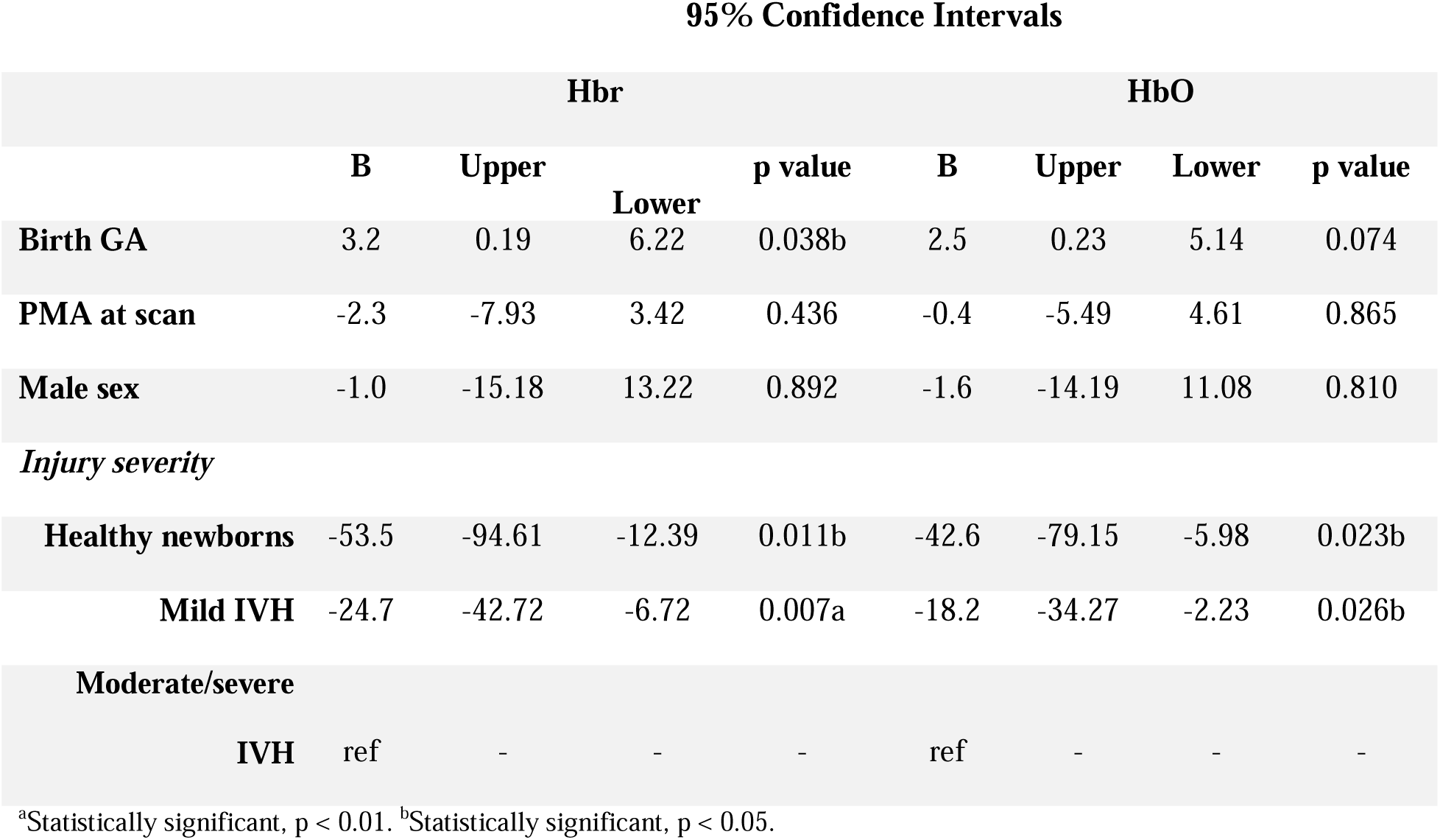
Results of a generalized linear model examining Hbr and HbO connectivity relative to IVH severity 95% Confidence Intervals.

## Discussion

In a prospective cohort of very preterm born infants with IVH, we examined the predictive utility of fNIRS connectivity in assessing brain health. As expected, RSFC maps were comparable between fMRI and fNIRS in the very preterm born infants with IVH. Findings indicate that fNIRS can be used to study cortical RSFC at the bedside. We further examined whether the severity of injury could be predicted by functional connectivity metrics. We found that for both Hbr and HbO, the connectivity values at TEA were increased in infants with moderate/severe IVH relative to healthy newborns and very preterm born infants with mild IVH. Overall, our results highlight the use of fNIRS as a bedside monitoring tool to examine brain-health metrics in very preterm born infants impacted by IVH.

In the first aim, we saw good correspondence of the RSFC maps (HbO vs fMRI and Hbr vs fMRI) between the two imaging modalities. Our findings are consistent with other studies. Duan et al., using fNIRS and fMRI data acquired from 21 adult subjects during resting state, demonstrated good correspondence between the two imaging modalities [17]. In another adult study, Sasai et al. also reported that fNIRS HbO and fMRI BOLD maps had a significant positive correlation for all brain regions investigated [23]. We were able to demonstrate similar results in our infant population. Second, except for the occipital region, the other regions (frontal, parietal and temporal) showed high similarity of RSFC maps between the two imaging modalities. The discrepancy in the occipital region could be because of measurement errors, including thick hair, poor contact between optodes and scalp, cap fit, which are common in fNIRS studies in infants [9],[13],[24]. In addition, all our bedside fNIRS measurements were carried out with the infants laying supine in their cot or caregivers’ arms which could explain the poor optode contact in the occipital region. Overall, our study adds to the body of literature showing that fNIRS indeed provides comparable RSFC measures to fMRI. This is especially important given the bedside availability of fNIRS for the vulnerable NICU population.

Filtering connections that are either negative- or low-weighted is a common practice for fNIRS RSFC research [25]. RSFC yielded from a series of sparsity levels were tested in previous studies, that demonstrated inconsistency among the results [26]–[28]. Recent studies have highlighted the necessity of choosing the proper sparsity value and have introduced various strategies [29],[30]. In the current work, we have addressed this issue by identifying a maximal similarity value for two imaging modalities (i.e., fNIRS and fMRI). For future research in fNIRS RSFC, this method may be impractical without access to another imaging modality such as fMRI. Yet, potentially this challenge can be overcome through scanning larger number of neonates, as this will aid in identifying a sparsity level that could be more universally appliable for neonatal RSFC studies.

Compared with their healthy counterparts, we found increased RSFC at TEA among infants with IVH, regardless of the severity. This was an unexpected finding, as most studies of very preterm infants with IVH have demonstrated reduced RSFC at TEA [14],[15],[31]. Reduced RSFC, especially in higher grades of IVH injury is attributed to disruptions to the periventricular white matter. However, some studies have also shown intact RSFC in infants with IVH [32]. The results from our study, while different from what is previously known in literature, could be explained by several plausible mechanisms. First, our study population was very preterm (mean GA 26.28 ±2.82), with IVH already evident within the first week of life and our findings represent the RSFC 10-12 weeks post IVH. Thereafter, during their NICU stay and with ongoing surveillance, depending on the IVH severity, some of the study participants underwent neurosurgical interventions to divert cerebrospinal fluid. In tandem with the above-mentioned, overtime the neonatal brain undergoes massive growth and reorganization, demonstrating a mature architecture by TEA [33],[34]. Moreover, some case studies have shown neural plasticity in infants with high grade IVH [35]. Hence, all these factors could explain our findings. Having said that, these questions are best addressed in longitudinal studies, correlating RSFC with neurodevelopmental outcomes. Disruptions in neonatal functional connectivity in children with perinatal brain injury have been associated with developmental outcomes, and in turn better characterization of these patterns is needed to improve early care practices [12].

Our study has several strengths, namely, both groups (infants with IVH and healthy controls) were recruited from and assessed at the same centre, using the same high-density fNIRS system and at similar postmenstrual ages. The above-mentioned measures ensure uniformity and eliminate potential bias. Second, compared to the clinical NIRS system that is now commonly used in most level III NICUs, high density fNIRS systems provide whole brain coverage and connectivity. Our study population (infants with IVH and healthy controls) was also well characterized. There are also some challenges and limitations that were associated with our study. Primarily, the sample size for our study population was small. This was due to difficulty acquiring excellent quality data, which is a challenge for fNIRS studies [24]. However, our eventual goal is to use fNIRS for clinical decision making in individual neonates at the bedside. Second, more males than females were recruited, due to lack of competitive enrolment. However, sex differences were not evident in any analyses. Despite the limitations, we believe that fNIRS offers promising avenues that can impact clinical care of infants with IVH. Larger prospective studies are needed to address these challenges.

## Conclusions

In a heterogenous cohort of very preterm born neonates with IVH who underwent fNIRS and fMRI we report comparable RSFC maps between the two modalities. Findings indicate, that in a small sample of neonates that bedside fNIRS can produce comparable results to that of fMRI. Secondly, fNIRS revealed distinct RSFC patterns between preterm infants with IVH at TEA and healthy infants. Larger prospective studies are needed to better characterize fNIRS-based functional connectivity changes over time and whether they are predictive of functional outcomes.

## Data Availability

All data produced in the present study are available upon reasonable request to the authors

## Data availability statement

The datasets generated and/or analysed during the current study are available upon reasonable request.

## Acknowledgments

We thank the families who participated in this study. We thank the staff from the Neonatal Intensive Care and Mother Baby Care Units at LHSC for their immense help during this study.

## Author contributions

L.M.N.K, L.T, T.A, A.K, M.M, S.R, H.V, P.M, P.C.M, S.d.R, S.B, L.T.A.R.d.M, M.T.J, K.S.L and E.G.D were involved in the study design, data acquisition design and execution of data analytic strategy, revision of the final version of this manuscript. L.M.N.K, L.T and E.G.D wrote the initial draft of the manuscript and contributed to the execution of the data analytic strategy. L.T and E.G.D analyzed the data. All authors approved the final manuscript as submitted and agree to be accountable for all aspects of the work.

## Consent statement

Consent was obtained from the guardians of the study participants.

## Statement of ethics

The study was approved by the Health Sciences Research Ethics Board at Western University. Informed consent was provided by the parents/caregivers of the infants enrolled in the study.

## References

1. Ballabh P. Pathogenesis and Prevention of Intraventricular Hemorrhage. Clin Perinatol 2014;41(1):47–67. Doi: 10.1016/j.clp.2013.09.007.

2. Siffel C, Kistler KD, Sarda SP. Global incidence of intraventricular hemorrhage among extremely preterm infants: A systematic literature review. J Perinat Med 2021;49(9):1017–26. Doi: 10.1515/JPM-2020-0331/ASSET/GRAPHIC/J_JPM-2020-0331_FIG_001.JPG.

3. McAllister JP, Guerra MM, Ruiz LC, Jimenez AJ, Dominguez-Pinos D, Sival D, et al. Ventricular zone disruption in human neonates with intraventricular hemorrhage. J Neuropathol Exp Neurol 2017;76(5):358–75. Doi: 10.1093/jnen/nlx017.

4. Papile LA, Burstein J, Burstein R, Koffler H. Incidence and evolution of subependymal and intraventricular hemorrhage: A study of infants with birth weights less than 1,500 gm. J Pediatr 1978;92(4):529–34. Doi: 10.1016/S0022-3476(78)80282-0.

5. Inder TE, de Vries LS, Ferriero DM, Grant PE, Ment LR, Miller SP, et al. Neuroimaging of the Preterm Brain: Review and Recommendations. J Pediatr 2021;237(0):276–287.e4. Doi: 10.1016/j.jpeds.2021.06.014.

6. Vohr BR. Neurodevelopmental outcomes of premature infants with intraventricular hemorrhage across a lifespan. Semin Perinatol 2022;46(5):151594. Doi: 10.1016/j.semperi.2022.151594.

7. Doria V, Beckmann CF, Arichi T, Merchant N, Groppo M, Turkheimer FE, et al. Emergence of resting state networks in the preterm human brain. Proc Natl Acad Sci U S A 2010;107(46):20015–20. Doi: 10.1073/pnas.1007921107.

8. Smyser CD, Snyder AZ, Shimony JS, Blazey TM, Inder TE, Neil JJ. Effects of White Matter Injury on Resting State fMRI Measures in Prematurely Born Infants. PLoS One 2013;8(7):e68098. Doi: 10.1371/journal.pone.0068098.

9. Wang Q, Zhu GP, Yi L, Cui XX, Wang H, Wei RY, et al. A Review of Functional Near-Infrared Spectroscopy Studies of Motor and Cognitive Function in Preterm Infants. Neurosci Bull 2020;36(3):321–9. Doi: 10.1007/S12264-019-00441-1/TABLES/1.

10. Triplett RL, Smyser CD. Neuroimaging of structural and functional connectivity in preterm infants with intraventricular hemorrhage. Semin Perinatol 2022;46(5):151593. Doi: 10.1016/j.semperi.2022.151593.

11. Duerden EG, Halani S, Ng K, Guo T, Foong J, Glass TJA, et al. White matter injury predicts disrupted functional connectivity and microstructure in very preterm born neonates. NeuroImage Clin 2019;21:101596. Doi: 10.1016/j.nicl.2018.11.006.

12. Linke AC, Wild C, Zubiaurre-Elorza L, Herzmann C, Duffy H, Han VK, et al. Disruption to functional networks in neonates with perinatal brain injury predicts motor skills at 8 months. Neuroimage (Amst) 2018;18:399. Doi: 10.1016/J.NICL.2018.02.002.

13. Scarapicchia V, Brown C, Mayo C, Gawryluk JR. Functional magnetic resonance imaging and functional near-infrared spectroscopy: Insights from combined recording studies. Front Hum Neurosci 2017;11(August):1–12. Doi: 10.3389/fnhum.2017.00419.

14. Argyropoulou MI, Xydis VG, Drougia A, Giantsouli AS, Giapros V, Astrakas LG. Structural and functional brain connectivity in moderate–late preterm infants with low-grade intraventricular hemorrhage. Neuroradiology 2022;64(1):197–204. Doi: 10.1007/S00234-021-02770-3/FIGURES/2.

15. Cha JH, Choi YH, Lee JM, Lee JY, Park HK, Kim J, et al. Altered structural brain networks at term-equivalent age in preterm infants with grade 1 intraventricular hemorrhage. Ital J Pediatr 2020;46(1):43. Doi: 10.1186/s13052-020-0796-6.

16. Kebaya LMN, Stubbs K, Lo M, Al-Saoud S, Karat B, St Lawrence K, et al. Three-dimensional cranial ultrasound and functional near-infrared spectroscopy for bedside monitoring of intraventricular hemorrhage in preterm neonates. Sci Reports 2023 131 2023;13(1):1–13. Doi: 10.1038/s41598-023-30743-4.

17. Duan L, Zhang YJ, Zhu CZ. Quantitative comparison of resting-state functional connectivity derived from fNIRS and fMRI: A simultaneous recording study. Neuroimage 2012;60(4):2008–18. Doi: 10.1016/j.neuroimage.2012.02.014.

18. Makropoulos A, Robinson EC, Schuh A, Wright R, Fitzgibbon S, Bozek J, et al. The Developing Human Connectome Project: a Minimal Processing Pipeline for Neonatal Cortical Surface Reconstruction Europe PMC Funders Group. Neuroimage 2018;173:88–112. Doi: 10.1101/125526.

19. Scholkmann F, Spichtig S, Muehlemann T, Wolf M. How to detect and reduce movement artifacts in near-infrared imaging using moving standard deviation and spline interpolation. Physiol Meas 2010;31(5):649–62. Doi: 10.1088/0967-3334/31/5/004.

20. Jahani S, Setarehdan SK, Boas DA, Yücel MA. Motion artifact detection and correction in functional near-infrared spectroscopy: a new hybrid method based on spline interpolation method and Savitzky-Golay filtering. Neurophotonics 2018;5(1):1. Doi: 10.1117/1.NPH.5.1.015003.

21. Kocsis L, Herman P, Eke A. The modified Beer-Lambert law revisited. Phys Med Biol 2006;51(5):N91–N98. Doi: 10.1088/0031-9155/51/5/N02.

22. Tantardini M, Ieva F, Tajoli L, Piccardi C. Comparing methods for comparing networks. Sci Rep 2019;9(1):1–19. Doi: 10.1038/s41598-019-53708-y.

23. Sasai S, Homae F, Watanabe H, Sasaki AT, Tanabe HC, Sadato N, et al. A NIRS-fMRI study of resting state network. Neuroimage 2012;63(1):179–93. Doi: 10.1016/j.neuroimage.2012.06.011.

24. Gallagher A, Wallois F, Obrig H. Functional near-infrared spectroscopy in pediatric clinical research: Different pathophysiologies and promising clinical applications. Neurophotonics 2023;10(02):023517. Doi: 10.1117/1.nph.10.2.023517.

25. Sasai S, Homae F, Watanabe H, Taga G. Frequency-specific functional connectivity in the brain during resting state revealed by NIRS. Neuroimage 2011;56(1):252–7. Doi: 10.1016/j.neuroimage.2010.12.075.

26. Nguyen T, Babawale O, Kim T, Jo HJ, Liu H, Kim JG. Exploring brain functional connectivity in rest and sleep states: a fNIRS study. Sci Reports 2018 81 2018;8(1):1–10. Doi: 10.1038/s41598-018-33439-2.

27. Einalou Z, Maghooli K, Setarehdan SK, Akin A. Graph theoretical approach to functional connectivity in prefrontal cortex via fNIRS. Neurophotonics 2017;4(04):041407. Doi: 10.1117/1.nph.4.4.041407.

28. Wang MY, Lu FM, Hu Z, Zhang J, Yuan Z. Optical mapping of prefrontal brain connectivity and activation during emotion anticipation. Behav Brain Res 2018;350:122–8. Doi: 10.1016/J.BBR.2018.04.051.

29. Chan YL, Ung WC, Lim LG, Lu CK, Kiguchi M, Tang TB. Automated Thresholding Method for fNIRS-Based Functional Connectivity Analysis: Validation with a Case Study on Alzheimer’s Disease. IEEE Trans Neural Syst Rehabil Eng 2020;28(8):1691–701. Doi: 10.1109/TNSRE.2020.3007589.

30. Dimitriadis SI, Salis C, Tarnanas I, Linden DE. Topological filtering of dynamic functional brain networks unfolds informative chronnectomics: A novel data-driven thresholding scheme based on orthogonal minimal spanning trees (OMSTs). Front Neuroinform 2017;11:238952. Doi: 10.3389/FNINF.2017.00028/BIBTEX.

31. Arichi T, Counsell SJ, Allievi AG, Chew AT, Martinez-Biarge M, Mondi V, et al. The effects of hemorrhagic parenchymal infarction on the establishment of sensori-motor structural and functional connectivity in early infancy. Neuroradiology 2014;56(11):985–94. Doi: 10.1007/S00234-014-1412-5/FIGURES/5.

32. Herzmann C, Zubiaurre-Elorza L, Wild CJ, Linke AC, Han VK, Lee DSC, et al. Using Functional Magnetic Resonance Imaging to Detect Preserved Function in a Preterm Infant with Brain Injury. J Pediatr 2017;189:213–217.e1. Doi: 10.1016/J.JPEDS.2017.06.063.

33. Fransson P, Skiöld B, Engström M, Hallberg B, Mosskin M, Åden U, et al. Spontaneous brain activity in the newborn brain during natural sleep-an fMRI study in infants born at full term. Pediatr Res 2009;66(3):301–5. Doi: 10.1203/PDR.0b013e3181b1bd84.

34. Eyre M, Fitzgibbon SP, Ciarrusta J, Cordero-Grande L, Price AN, Poppe T, et al. The Developing Human Connectome Project: Typical and disrupted perinatal functional connectivity. Brain 2021;144(7):2199–213. Doi: 10.1093/brain/awab118.

35. Guzzetta A, Fiori S, Scelfo D, Conti E, Bancale A. Reorganization of visual fields after periventricular haemorrhagic infarction: Potentials and limitations. Dev Med Child Neurol 2013;55(SUPPL.4):23–6. Doi: 10.1111/dmcn.12302.

